# Tolerability, safety and immunogenicity of intradermal delivery of a fractional dose mRNA-1273 SARS-CoV-2 vaccine in healthy adults as a dose sparing strategy

**DOI:** 10.1101/2021.07.27.21261116

**Authors:** Geert V.T. Roozen, Margaretha L.M. Prins, Rob van Binnendijk, Gerco den Hartog, Vincent P. Kuiper, Corine Prins, Jacqueline J. Janse, Annelieke C. Kruithof, Mariet C.W. Feltkamp, Marjan Kuijer, Frits R. Rosendaal, Meta Roestenberg, Leo G. Visser, Anna H.E. Roukens

**Affiliations:** Department of Parasitology, Leiden University Medical Center, Leiden, The Netherlands; Department of Infectious Diseases, Leiden University Medical Center, Leiden, The Netherlands; Department of Immune Surveillance, National Institute for Public Health and the Environment, The Netherlands; Centre for Human Drug Research, Leiden, The Netherlands; Department of Medical Microbiology, Leiden University Medical Center, Leiden, The Netherlands; Department of Clinical Epidemiology, Leiden University Medical Center, Leiden, The Netherlands

## Abstract

**Background:** There is an urgent need for fair and equitable access to safe and effective vaccines to end the COVID-19 pandemic. Shortages in reagents and vaccines are a major challenge, as well as limited knowledge on dose response relationship with mRNA COVID-19 vaccines. We explored intradermal fractional dose administration of a mRNA SARS-CoV-2/COVID-19 vaccine as a potential dose-sparing strategy.

**Methods:** We conducted a proof-of-concept, dose-escalation, open-label, randomised-controlled vaccine trial (IDSCOVA) in healthy adults aged 18-30 years. To test initial safety, ten participants received 10 µg mRNA-1273 vaccine through intradermal injection at day 1 and 29. Following a favourable safety review, thirty participants were 1:1 randomised to receive 20 µg mRNA-1273 either intradermally or intramuscularly. The primary endpoint was tolerability and safety. The secondary endpoint was seroconversion and specific IgG concentration against SARS-CoV-2 spike S1 and Receptor Binding Domain (RBD) after the second dose at day 43. We compared results to two historical cohorts of non-hospitalised COVID-19 patients and vaccinated individuals.

**Findings:** Thirty-eight of forty included participants (median age 25 years) completed the study. There were no serious adverse events. Self-reported local adverse reactions after intradermal delivery were mild, both in the 10 µg and the 20 µg group. In the higher dose group, systemic adverse reactions were more common, but still well tolerated. All 38 participants mounted substantially higher IgG-anti-S1 and IgG-anti-RBD concentrations at day 43 than COVID-19 controls. At day 43, anti-S1 (95% CI) was 1,696 (1,309-2,198) BAU/mL for the 10 µg intradermal group, 1,406 (953·5-2,074) BAU/mL for the 20 µg intramuscular group and 2,057 (1,421-2,975) BAU/mL for the 20 µg intradermal group. Anti-S1 was 107·2 (63-182·2) BAU/mL for the convalescent plasma control group and 1,558 (547·8-4,433) BAU/mL for the individuals vaccinated with 100 µg mRNA-1273.

**Interpretation:** Intradermal administration of 10 µg and 20 µg mRNA-1273 vaccine was well tolerated and safe, and resulted in a robust antibody response. Intradermal vaccination has the potential to be deployed for vaccine dose-sparing.

**Funding:** The trial was supported by crowdfunding (Wake Up to Corona).

## Introduction

The severe acute respiratory syndrome coronavirus-2 (SARS-CoV-2) has caused a worldwide pandemic since December 2019. The virus has now affected over 190 million people.^1^ SARS-CoV-2 is still spreading in many parts of the world resulting in the rise of multiple variants of concern through adaptive deletions and mutations.^2^ These variants are generally characterised by an increased transmissibility and decreased susceptibility to neutralising antibodies, making a fast, fair and equitable access to safe and effective COVID-19 vaccines for all countries of the world of utmost importance to protect the most vulnerable and to halt the devasting global morbidity and mortality.^3^

Currently, five vaccines are listed by the World Health Organization (WHO) for emergency use.^4^ These vaccines are limited in supply, especially in low- and middle-income countries, leading to substantial morbidity and mortality.^5^ A large unvaccinated population continues to pose a threat of a continuous emergence of new SARS-CoV-2 variants that may be more infectious, more malignant and more resistant to vaccines than current strains.^6^ Despite the COVID-19 Vaccines Global Access (COVAX) Facility initiated by the WHO to provide vaccine access for low-income countries, probably 80% of the vaccine needs of participating countries will not be met soon.^7^ Exploring dose-sparing techniques could therefore provide the solution to immunise more people with the same vaccine stockpile.

The intramuscular injection (IM) is the standard inoculation route of vaccines. However, the papillary dermis contains a higher density of antigen presenting dendritic cells (APC) than muscle tissue, which makes it an obvious compartment to administer vaccine antigens into.^8^ In addition, the dermal lymphatic system is organised into several plexus systems, which aids efficient transport of vaccine antigen and APCs to the draining regional lymph nodes where further activation of B- and T-lymphocytes takes place and a protective immune response is mounted.^9^ As a consequence, the intradermal delivery (ID) of a fractional vaccine dose (for example 1/10^th^ or 1/5^th^ of the standard dose) introduced directly into the papillary dermis can be as effective as the IM administration of the standard dose. This has already been demonstrated for rabies, yellow fever, inactivated polio and seasonal influenza vaccine.^10^

Several preclinical studies support the efficacy of the intradermal route for mRNA lipid-nanoparticle (LNP)-based vaccines. In mice, it has been demonstrated that the total amount of protein produced was higher after ID delivery than IM delivery at the lowest dose of mRNA-LNP. At higher doses, saturation was reached which favoured IM delivery, showing that especially for lower mRNA doses ID delivery is the preferred route.^11^ In Rhesus macaques intradermal H10-mRNA-LNP influenza vaccine delivery resulted in higher antibody titers after the second dose, than IM delivery.^12,13^

In humans, the safety and immunogenicity of the ID administration of H10N8 mRNA-LNP influenza vaccine has been evaluated.^14^ The 25 µg ID dose induced hemagglutination inhibition titers >1:40 in 65% of participants compared with 34% of participants who received the same dose IM, confirming the high immunogenicity of the ID route at low dose. The most frequently reported adverse reactions after ID injection were: pain (82%), erythema (76%) and swelling (38%) at the injection site, and fatigue (38%), headache (26%), myalgia (14%), nausea (9%) and arthralgia (3%). No severe adverse reactions were reported. Since the ID vaccination route was associated with higher rates of solicited local reactions, the sponsor elected to discontinue the enrollment for ID administration in the study.

Given the urgent need to increase the availability of COVID-19 vaccines, we investigated the safety and tolerability of mRNA-1273 vaccine delivered intradermally. This lipid-nanoparticle (LNP)- encapsulated mRNA-1273 vaccine was developed by Moderna and the Vaccine Research Center (VRC) of the National Institute of Allergy and Infectious Disease (NIAID). The mRNA vaccine is encoding for the prefusion-stabilized form of the SARS-CoV-2 spike protein (S-protein) and has a favourable thermostability. Since all mRNA vaccines are vulnerable to degradation at room temperature, they must be stored and transported frozen, via a cold supply chain.^15^ However, mRNA-1273 vaccine can be maintained for 30 days at 2-8 °C, which increases feasibility of use.^15,16^ The efficacy of two IM doses of 100 µg mRNA-1273 vaccine was recently demonstrated to be 94% for the prevention of COVID-19 illness as compared with placebo.^17^

Here, we report the safety and immunogenicity of the intradermal delivery of 10 µg and 20 µg mRNA-1273.

## Methods

### Study design and participants

This study was designed as a proof-of-concept, dose-escalation, open-label, randomized-controlled vaccine trial (IDSCOVA) conducted at the Leiden University Medical Center in collaboration with the Centre for Human Drug Research in the Netherlands. In the first part the tolerability, safety and immunogenicity of intradermal administration of two doses of 10 µg mRNA-1273 at 28-days interval was established. Following safety review by an independent Data Safety Monitoring Board, the tolerability, safety and immunogenicity of two doses of 20 µg mRNA-1273 through ID or IM route was established in the second part.

Eligible participants were healthy adults aged 18 to 30 years. Participants were screened for SARS-CoV-2 infection by serology [SARS-CoV-2 anti-N IgG antibodies (Liaison by Diasorin, Sallugia, Italy) and SARS-CoV-2 PCR of a mid-turbinate/throat swab and excluded when positive. Persons were excluded when one of the following criteria were met: having received a COVID-19 vaccine, at risk for severe COVID-19 disease, history of severe adverse or allergic reactions to previous vaccinations or any component of the study intervention, known or suspected immunodeficiency, history of autoimmune disease or active autoimmune disease requiring therapeutic intervention, use of systemic or topical corticosteroids, bleeding diathesis, pregnancy, breastfeeding or planned pregnancy within four weeks after final injection.

The trial was performed in accordance with the ethical principles of the Declaration of Helsinki and Good Clinical Practice guidelines developed by the International Harmonisation Task Force. The protocol was approved by the Medical Ethical Committee Leiden, Den Haag, Delft (NL 76702.058.21) and registered in the Netherlands Trial Register (NTR) (https://www.trialregister.nl/trial/9275). Safety was reviewed by an independent Data Safety and Monitoring Board. All participants provided written informed consent. The vaccine manufacturer was not involved in this trial.

For comparison of immunogenicity, antibody titers were compared with two previously collected cohorts of PIENTER-Corona. PIENTER-Corona is an ongoing nationwide study including all age groups of the Dutch general population, with serum samples collected at regular time intervals irrespective of the moment of infection or vaccination. For this study, convalescent sera (n=23) of PCR-confirmed patients were selected that had been collected between two weeks and two months after symptom onsets from individuals aged 18-45 years with mild to moderate COVID-19 (Table S3). A second set of 20 samples were selected of vaccinated individuals without an infection history, aged 18-65 years, collected between 14 and 45 days after receiving the second IM vaccination with 100 µg mRNA-1273 vaccine. The PIENTER-Corona study was approved by the medical ethical committee MED-U, Nieuwegein, the Netherlands and registered in NTR (https://www.trialregister.nl/trial/8473).

### Randomisation and masking

The first part was open label. In the second part, a 1:1 randomisation was performed with sealed envelopes. Investigators and participants were not blinded; laboratory personnel was blinded for the study groups.

### Procedures

In part one, ten participants received 10 µg mRNA through the ID route. The vaccine was prepared from the standard mRNA-1273 vaccine with a concentration of 100 µg mRNA per 0·5mL. We administered 0·05 mL ID injection in the deltoid region using a two-dose regimen with doses on day 1 and day 29. In part two, thirty participants were randomised 1:1 to receive either 20 µg mRNA-1273 ID or IM with also doses on day 1 and day 29. With the standard ID injection technique, the needle was inserted at a 5-to-15 degree angle and advanced through the epidermis for approximately 3 mm to ensure that the entire bevel was covered by the skin. After injection a tense, pale wheal of 6 to 10 mm in diameter will appear over the needle bevel. The second vaccination was administered at the contralateral site to distinguish between local adverse reactions after the first and second vaccination. After each injection, the wheal size was measured, quantified in mm as wheal diameter across two perpendicular axes. Participants were observed for 30 minutes after vaccination to monitor acute reactions. For both part 1 and 2, the dose-escalation design specified that for each dose three sentinel participants were enrolled and monitored first. If none of the pre-specified stopping criteria were met in the first 48 hours after vaccination of the first three sentinel participants, the remaining participants were enrolled (Supplement).

### Monitoring of tolerability and safety

Follow-up visits by phone were scheduled on day 2, 3, 4, 8, 15, 30, 31, 32 and 57 to assess safety outcomes. Physical follow-up visits were scheduled on day 36 and 43, i.e., seven and 14 days after the second vaccination, respectively. At each physical visit SARS-CoV-2 infection was excluded by serology and PCR. After each vaccination, participants recorded their temperature and adverse events (AEs) daily in a diary for 14 days (Supplement). Solicited AEs included nature and severity of local reactions at the injection site and regional lymph nodes, and nature and severity of systemic reactions. All local and systemic reactions were graded according to a standard grading scale (Table S1-S3). Participants were instructed not to use analgesics or antipyretics routinely before or after the vaccinations. Any use of antipyretic or pain medication that occurred in the 14 days after administration of the study intervention was registered.

### Safety assessments

The primary objective of this study was to evaluate the safety, assessed by the nature, frequency and severity of solicited and unsolicited local and systemic adverse reactions and the use of antipyretics and analgesics. A secondary outcome was the diameter of the wheal in mm after intradermal injection, as a quality indicator of the intradermal vaccination technique.

### Immunogenicity

Serum samples were collected at screening and at days 29, 36 and 43. All sera were tested for the presence of specific IgG antibodies against the nucleoprotein, Spike S1 protein and the receptor binding domain (RBD), located in the S1 subunit using a bead-based multiplex immunoassay (MIA) based on Luminex technology, as previously described.^18,19^ Antibody concentrations were interpolated from a 5-parameter fit of a serum pool calibrated against the WHO international reference (NIBSC, no 20/136) and reported in binding-antibody units (BAU/mL, as recently reported.^20^ Cutoffs for seropositivity were set to 10·08 BAU/mL for S1 and 30·29 for RBD with a specificity of 99·7% and 99·0% and a sensitivity of 90·9% and 91·6% for S1 and RBD respectively.

In addition, we used MIA for detection of anti-nucleocapsid (anti-N) antibodies in parallel to assess any occurrence of SARS-CoV-2 infection following vaccination. Screening at inclusion of participants for presence of anti-N IgG was performed with the Liaison assay, as this is the in-house test available and provided a rapid result.

### Statistical analysis

The sample size for this study was based on the probability of observing at least one AE for a given true event rate of a particular AE, for a specific sample size. If the true AE rate is 10%, then there is 65% probability of observing at least one AE in part one with 10 participants and 79% in part two with 15 participants. All participants who received at least one dose of the study intervention were included for the safety analysis. Descriptive summary statistics included numbers and percentages with the indicated endpoint and the associated Clopper Pearson 95% Confidence Intervals (CI). All AEs were categorized according to the International Classification of Diseases-10 (ICD10) terms. Wheal diameter was reported as mean with standard deviation and range. Immunogenicity data were analysed using summary descriptive statistics and included all eligible randomised participants with at least one valid immunogenicity result. SARS-CoV-2-specific antibody concentrations against S1 and RBD were expressed as geometric mean concentrations (GMCs) and 2-sided 95% geometric CI. Differences between groups were evaluated using the non-parametric Kruskal-Wallis test reporting adjusted p-values. All analyses were conducted using IBM SPSS Statistics for Windows, version 25.0. Armonk, New York: IBM Corp. Graphs were made by using GraphPad Prism version 9.0.1 for Windows, GraphPad software, San Diego, California.

## Results

### Study population

Between April 15^th^ and May 3^rd^, 2021, ten participants were included for part one, and thirty were included for part 2. After the first vaccination, one participant withdrew as she was offered a regular COVID-19 vaccination by her employer and one participant withdrew due to personal reasons (Figure 1). The characteristics of included participants are given in Table 1.

**Table 1.** Characteristics of participants at inclusion. General characteristics of the participants at inclusion. SD = Standard Deviation. BMI = Body Mass Index. ID = Intradermal. IM = Intramuscular.

|  | 10 µg ID<br>(n=10) | 20 µg ID<br>(n=15) | 20 µg IM<br>(n=15) |
| --- | --- | --- | --- |
| Female, n (%) | 4 (40) | 9 (60) | 7 (47) |
| Age, years |  |  |  |
| Mean (SD) | 24 (4) | 26 (3) | 24 (3) |
| Range | 18-28 | 19-29 | 19-30 |
| BMI, kg/m<sup>2</sup> |  |  |  |
| Mean (SD) | 24 (4) | 26 (3) | 24 (3) |
| Range | 20-25 | 20-30 | 19-30 |

**Figure 1.**
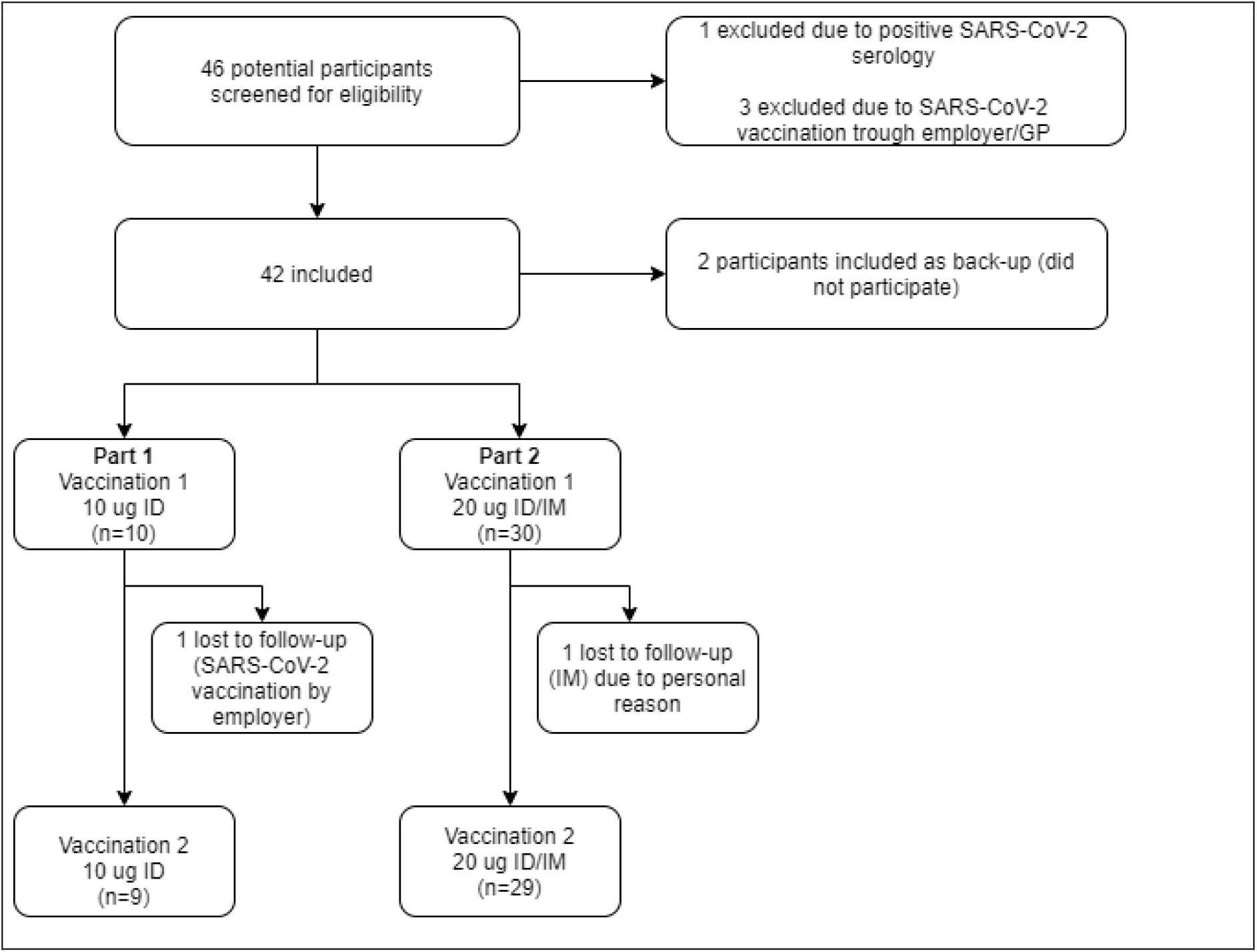
Flow-chart of inclusions. Flow-chart of inclusions in part 1 and 2. ID = Intradermal, IM = Intramuscular, GP = General Practitioner

### Safety

There were no serious adverse events (SAE) and none of the pre-specified study stopping rules were met. An overview of vaccine-related adverse reactions is shown in Figure 2. No acute adverse reactions occurred in the first 30 minutes after vaccine administration. In total, four participant used antipyretic medication to treat vaccine related AE after the first vaccination and 4 participants used antipyretic medication after the second vaccination (Table S4). Most AEs lasted 1 or 2 days on average with no AE lasting longer than 7 days, with an exception to hyperpigmentation (Table S5), which lasted up to 44 days in one participant (Figure S1A).

**Figure 2.**
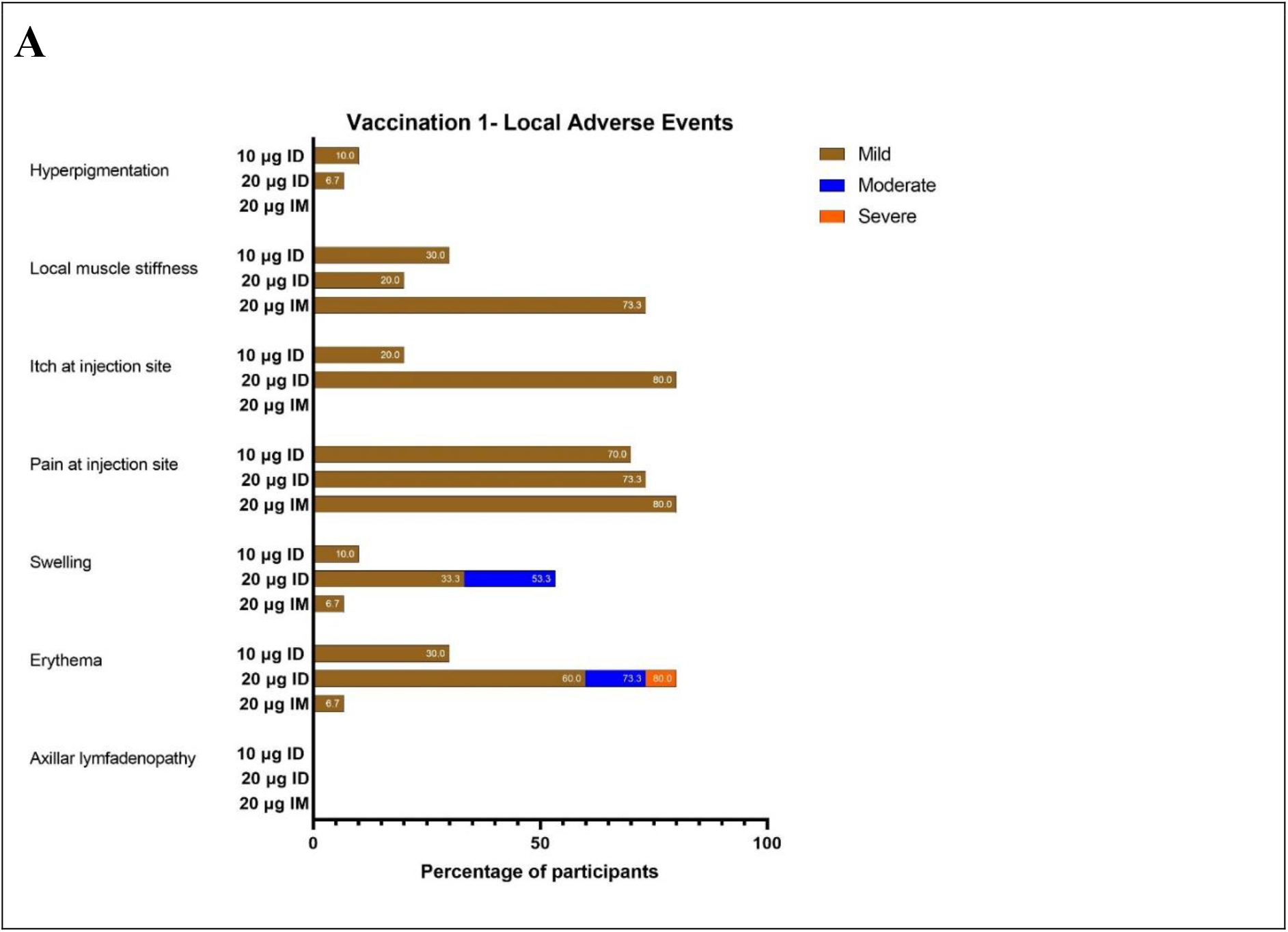
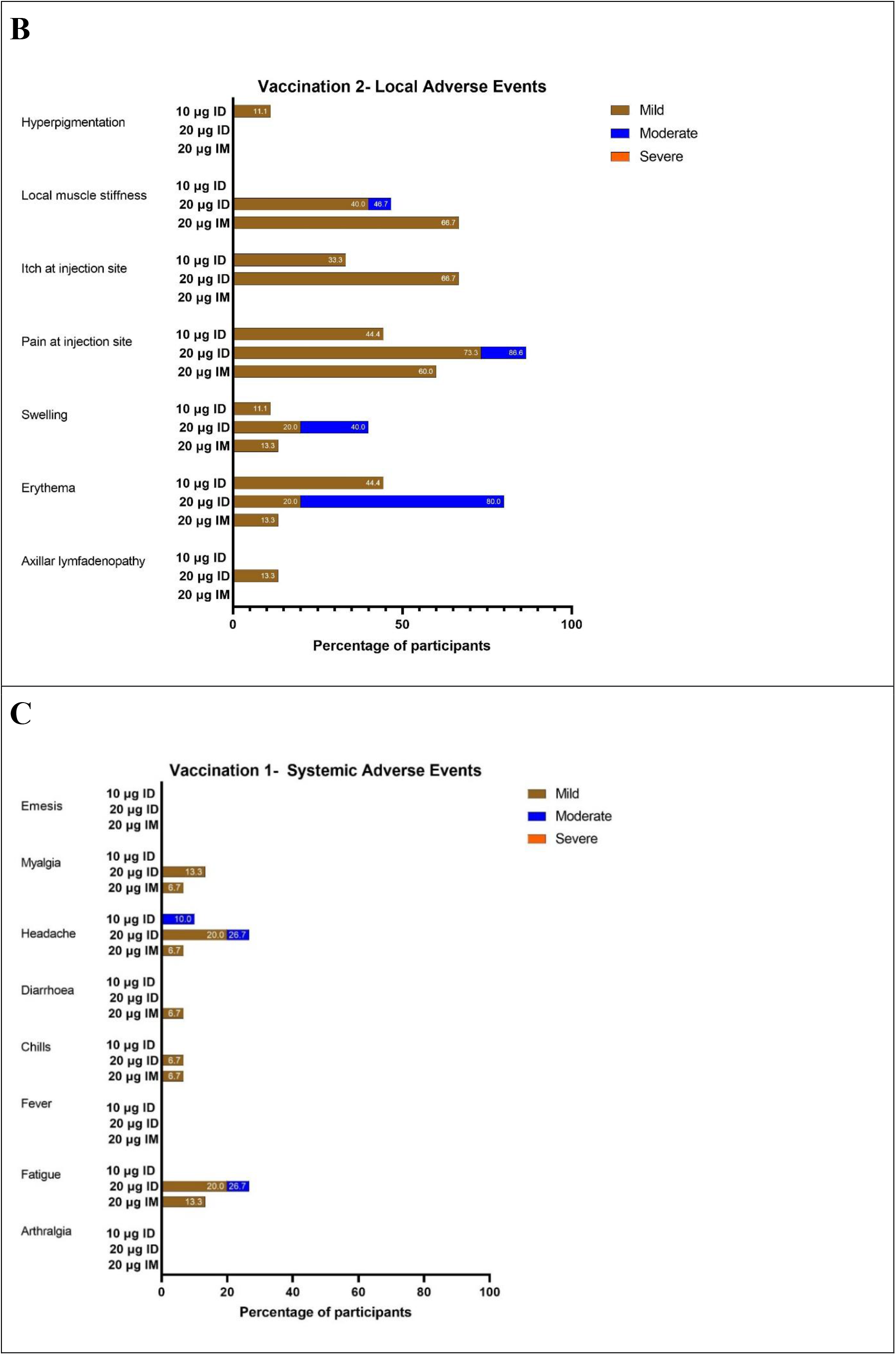
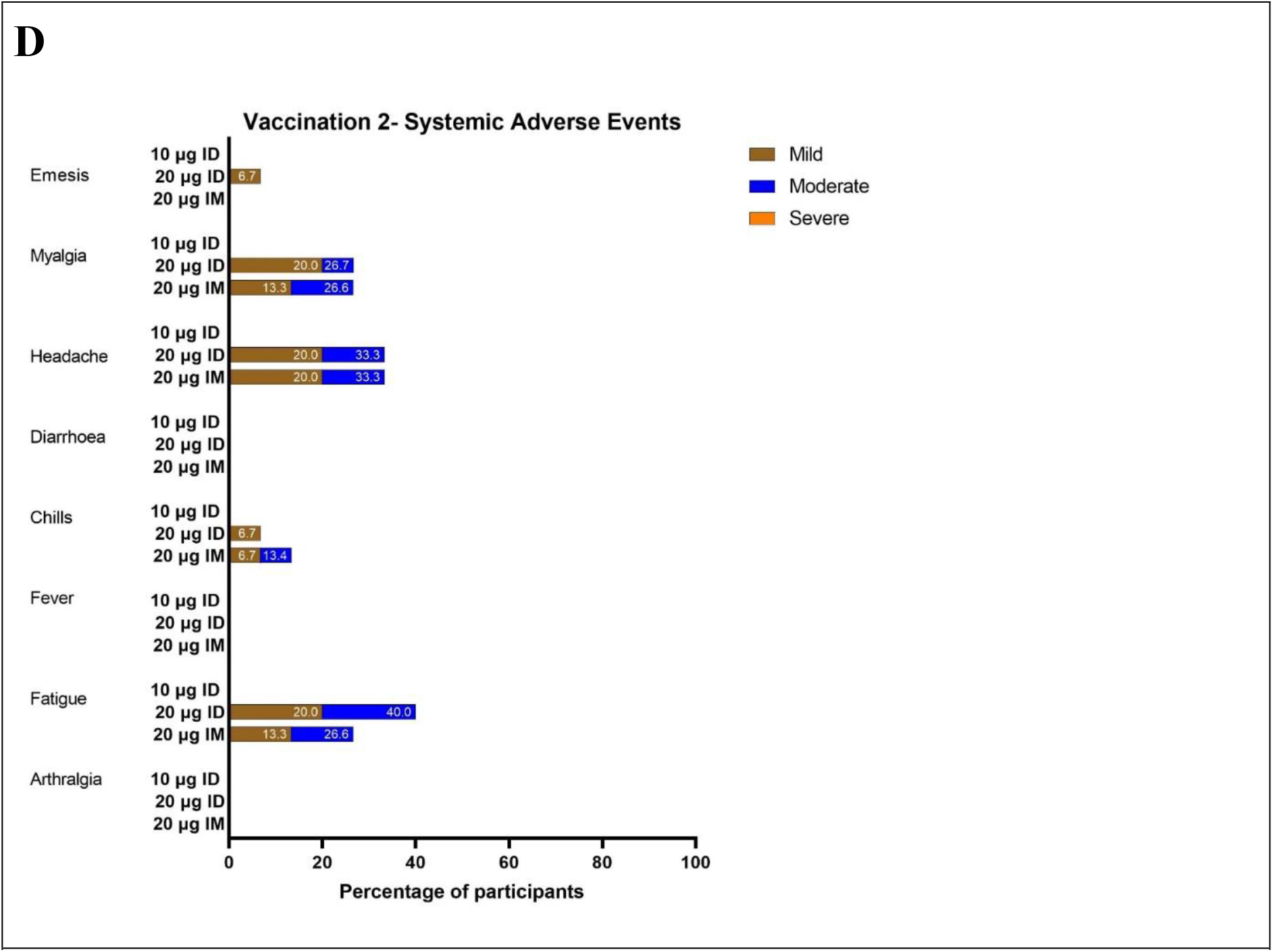
Local and systemic adverse events related to the vaccination. Local adverse (A and B) and systemic (C and D) events related to the vaccination in part 1 and part 2, subdivided in mild, moderate or severe. All adverse events possibly, probably or definitely related to the vaccination in the following 28 days after the first (A) and second (B) vaccine administration. ID = Intradermal, IM = Intramuscular. Numbers in bars represent percentage of participants reporting this adverse event.

### Local AEs

After the first vaccination in the 10 µg ID group, the most commonly reported local adverse reaction was pain at the injection site (70%). After the second vaccination, nine participants experienced local reactions, all reported as mild. The most prevalent AEs were erythema (44·4%), pain (44·4%), and itch (33%) at the injection site.

In the 20 µg groups, ID vaccination elicited more local AEs than IM vaccination.

After the first vaccination, most common local AEs in the 20 µg ID group were itch (80%) and erythema (80%). One participant in this group reported severe erythema of more than 10 cm in diameter and moderate swelling. This lasted for about six days, was self-limiting and well tolerated (Figure S1C). Most common local AEs in the 20 µg IM group was pain at the injection site (80%).

In both the 20 µg ID and IM group, local AEs were less common after the second vaccination. Local pain (86·6%) and erythema (80%) were most reported after the second 20 µg ID vaccination. In the 20 µg IM group, local muscle stiffness (68·7%) and local pain (60%) were most reported after the second vaccination.

Information on average diameter of swelling and erythema is shown in Figure S5. The ID administrations elicited more local swelling and erythema than the IM administration, and 20 µg ID more than 10 µg ID. In both ID groups, there was a reoccurrence of erythema and swelling observed eight to ten days after the first vaccination. In all groups, the second vaccination caused more erythema and swelling than the first vaccination.

### Systemic AEs

In the 10 µg ID group, the only reported systemic AE was moderate headache in one participant (10%) after the first vaccination.

In both the 20 µg ID and IM group, the second vaccination elicited more systemic AEs than the first.

In the 20 µg ID group, the most reported systemic AEs after the first and second vaccination were headache (26·7% and 33·3%, respectively) and fatigue (26·7%, 40·0%, respectively). In the 20 µg IM group, the most reported systemic AE was fatigue (13·3%) after the first vaccination and headache (33·3%) after the second vaccination.

### Wheal diameter after intradermal vaccination

The average wheal size after the first and second intradermal vaccination with 10 µg (0·05 mL) was 8 mm (SD 1; range 6 – 9 mm) and 7 mm (SD 1; range 7 – 8 mm), respectively. For intradermal vaccination with 20 µg (0·10 mL), average wheal size was 8 mm (SD 1; range 6 – 10 mm) and 10 mm (SD 1; range 8 – 10 mm), respectively for the first and second vaccination.

### Immunogenicity

All participants seroconverted after the first vaccine dose (Figure S3). All participants remained anti-Nucleoprotein IgG negative during the study (Figure 3c), indicating that their anti-SARS-CoV-2 response was elicited by the mRNA-1273 vaccine and not through natural infection.

**Figure 3.**
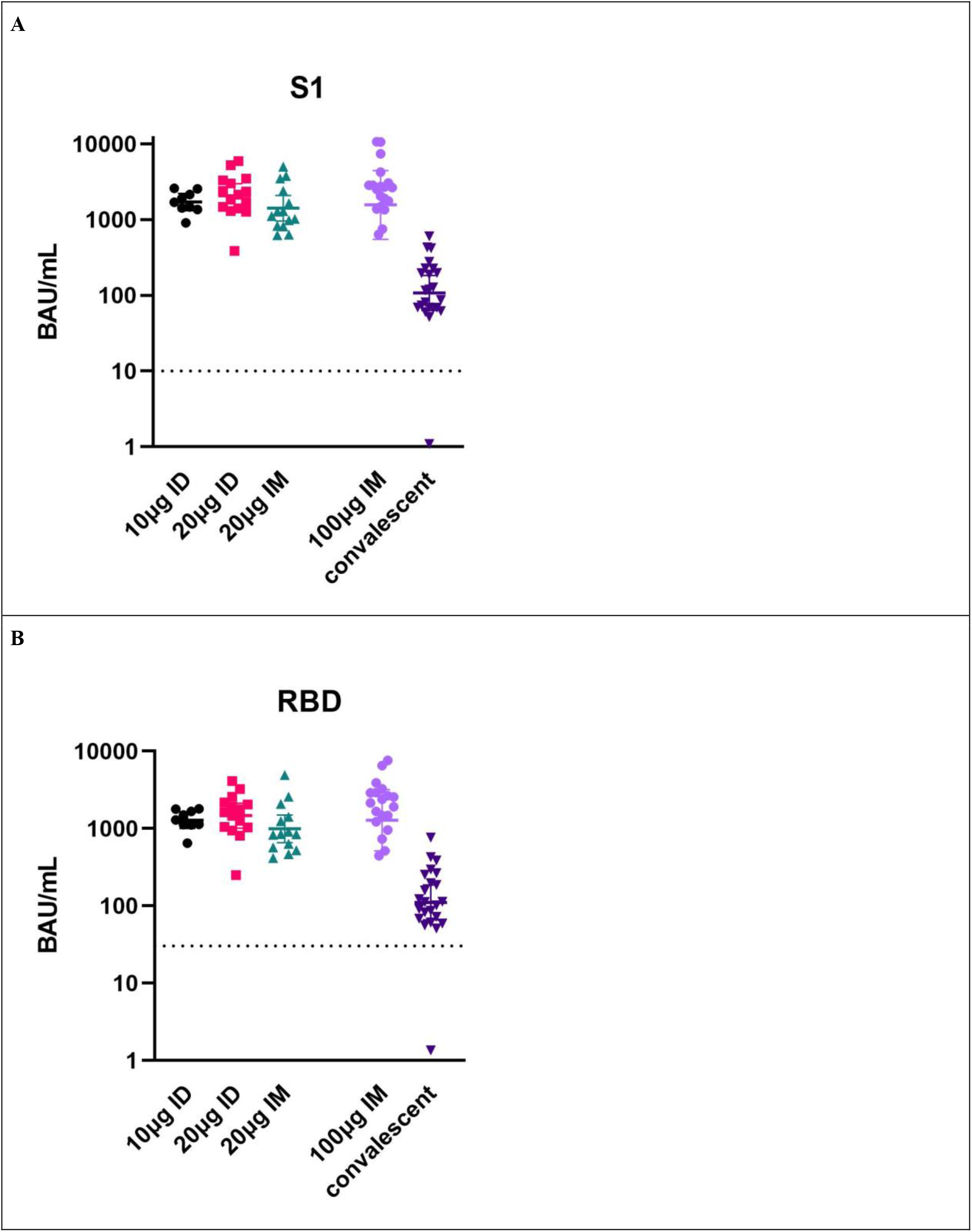
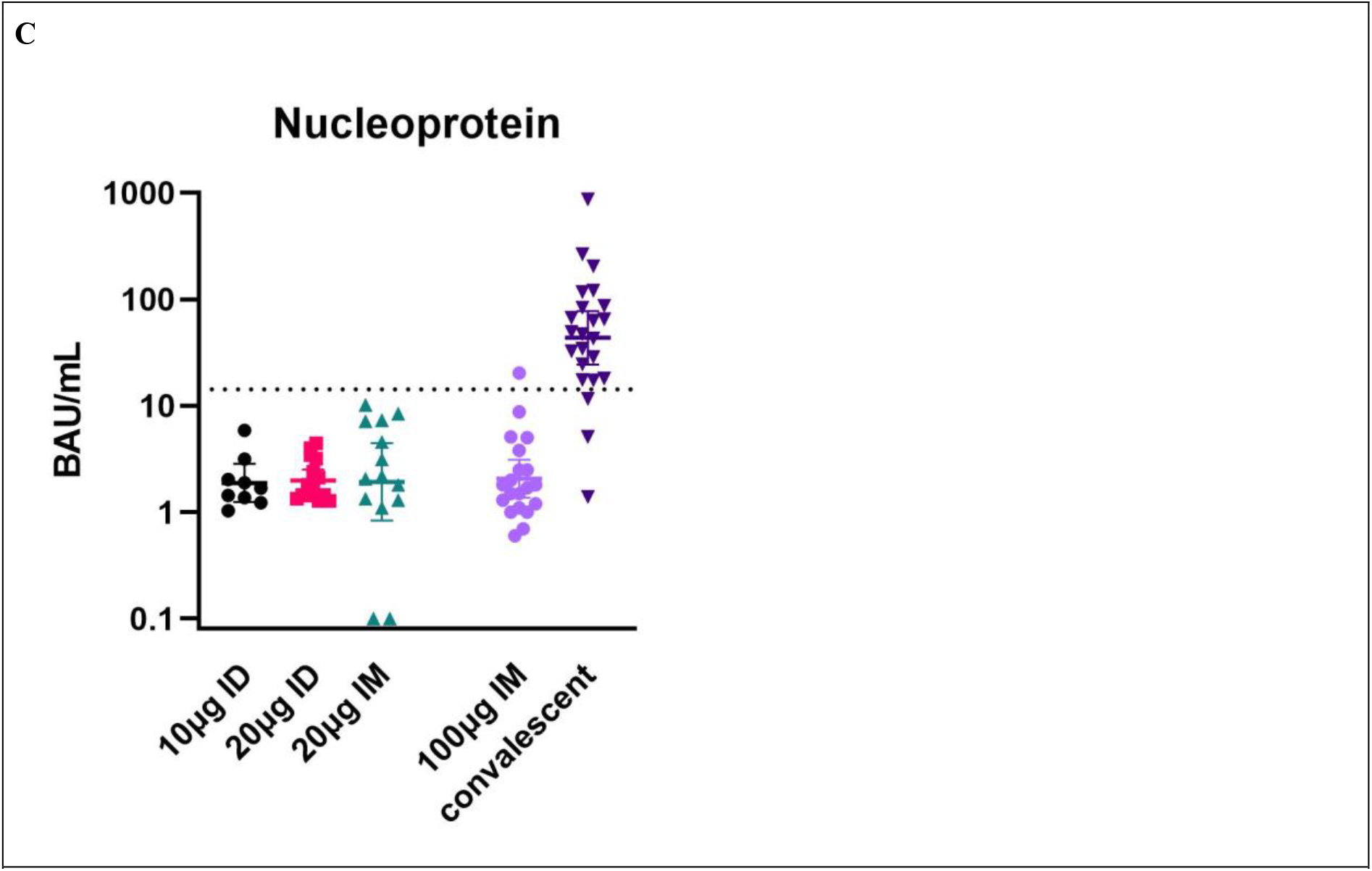
Anti-SARS-CoV-2 antibody concentrations. Anti-SARS-CoV-2 antibody concentrations by bead-based immunoassay (MIA) a) Anti-Spike S1; b) Anti-receptor binding domain IgG and c) Anti-Nucleoprotein IgG concentrations reported in binding antibody units/mL at day 43 in part 1 (10 μg ID) and part 2 (20 μg ID or IM). Each symbol represents a sample from an individual participant. Convalescent antibody concentrations were measured in symptomatic, non-hospitalised patients. Samples from persons vaccinated with 100 µg mRNA-1273 vaccine IM were also selected for reference. Values above dotted line are positive results. Concentrations of S1 and RBD-IgG levels were significantly higher in ID vaccinated persons compared to levels observed in convalescent sera (p-values < 0.0008).

For part 1, anti-S1 IgG titers increased rapidly after the first and second vaccination in all participants and amounted to 1,696 [95% CI 1,309-2,198] BAU/mL on day 43 (Table 2). Similar results were found for antibodies to the RBD: anti-RBD IgG titers were 1,286 [95% CI 1,004-1,648] BAU/mL on day 43 (Table 3).

**Table 2.** IgG antibodies against Spike S1 protein of SARS CoV-2.

|  | **Part 1**<br>**10 µg ID**<br>**(n=9)** | **Part 2**<br>**20 µg ID**<br>**(n=15)** | **Part 2**<br>**20 µg IM**<br>**(n=14)** |
| --- | --- | --- | --- |
| Day 1<br>(Vaccination 1) | 0.8<br>(0.430-1.49) | 0.32<br>(0.14-0.68) | 0.25<br>(0.162-0.415) |
| Day 29<br>(Vaccination 2) | 337.1<br>(237.1-479.2) | 405.6<br>(263.7-623.8) | 233<br>(142.2-381.8) |
| Day 36 | 813.8<br>(590.3-1,171.9) | 972.8<br>(672.8-1,409) | 612.0<br>(342.7-1,093) |
| Day 43 | 1,696<br>(1,308.9-2,197.9) | 2,057<br>(1,421-2,975) | 1,406<br>(953.5-2,074) |

Geometric mean concentrations of anti-S1 antibodies (95% CI) in BAU/mL. BAU = Binding Antibody Units, CI = Confidence Intervals. Cut off for seropositivity is 10.08 BAU/mL.

**Table 3.** IgG antibodies against RBD of SARS CoV-2.

|  | **Part 1**<br>**10 µg ID**<br>**(n=9)** | **Part 2**<br>**20 µg ID**<br>**(n=15)** | **Part 2**<br>**20 µg IM**<br>**(n=14)** |
| --- | --- | --- | --- |
| Day 1<br>(Vaccination 1) | 1.6 (0.9-3.3) | 1 (0.6-1.8) | 0.8 (0.6-1.1) |
| Day 29<br>(Vaccination 2) | 201.8 (141.8-287.3) | 257 (166.8-396) | 134.3 (80.9-222.8) |
| Day 36 | 580 (413.9-812.6) | 631.9 (444.7-898.1) | 367.4 (203.6-663.2) |
| Day 43 | 1,286 (1,003-1,648) | 1,471 (1,013-2,137) | 993.5 (659.5-1,497) |

Geometric mean concentrations of anti-RBD antibodies (95% CI) in BAU/mL. BAU = Binding Antibody Units, CI = Confidence Intervals. Cut off for seropositivity is 10.08 BAU/mL.

For part 2, anti-S1 IgG titers were 2,057 BAU/mL [95% CI 1,421-2,975] on day 43 in the ID group, and 1,406 BAU/mL [95% CI 953·3-2,074] in the IM group (Table 2). Similar results were found for the antibodies to the RBD (Table 3)

Concentrations of S1 and RBD-specific IgG were 14 to 20-fold higher in ID-vaccinated individuals than in convalescent sera from symptomatic, non-hospitalised patients. In these patients Anti-S1 and anti-RBD IgG titers at day 49 were 107·2 [95% CI 63-182·2] and 110·9 [95% CI 65·8-187·1], respectively (Figure 3a and 3b). IgG concentrations after 10 or 20 μg ID and 20 μg IM were similar to those observed in individuals of the general population after receiving 100 μg IM (Figure 3a and 3b). In the latter group anti-S1 and anti-RBD titers in these patients at day 49 were 1,558;95% CI 547·8-4433] and 1286 [95% CI 516-3205], respectively.

## Discussion

We report results of a proof of concept, open-label, dose-escalation vaccine trial, and show that intradermal delivery of a two-dose regimen of 10 µg or 20 µg fractional dose of mRNA-1273 SARS-CoV-2 vaccine in healthy adults was safe, well tolerated and immunogenic. These findings support further exploration of the intradermal delivery of a fractional dose of mRNA-1273 vaccine as a dose-sparing strategy. Fractional dose intradermal vaccination has previously been investigated as a dose-sparing strategy for several vaccines (e.g. influenza, rabies, hepatitis A and B, measles, DTP),^9^ but not for a registered COVID-19 vaccine.

The most commonly reported adverse reactions were short-lasting and consisted of mild pain, itching, erythema and swelling at the injection site. These local reactions are similar to those reported after intradermal vaccination with other vaccines.^10^ Delayed injection-site reactions, with an onset of ten days or more after injection, were uncommon.

In this study, we showed that a two-dose regimen of 10 µg or 20 µg mRNA SARS-CoV-2 vaccine, administered intradermally, induced a homogeneous rapid and robust increase in anti-S1 and anti-RBD antibodies by day 43 in all participants. These titers were considerably higher (14 to 20-fold) than antibody concentrations in convalescent samples. Post-vaccination titers were in the same range as samples from individuals vaccinated with two doses of 100 µg mRNA-1273, suggesting that a dose-sparing strategy using 1/10^th^ or 1/5^th^ dose of mRNA-vaccine provided sufficient protective immunity. The reduced dosage did not result in any primary vaccine failure among the twenty-four volunteers who received the vaccine at reduced dosage.

The relationship between specific IgG concentrations and the capacity to neutralise SARS-CoV-2 is currently under investigation. A first indication of the degree of neutralising capacity of antibodies induced by ID administration as presented in this study can be derived from a recent study showing the relationship between vaccine efficacy and virus neutralisation titers.^21^ Sera from mRNA-1273 vaccinated individuals had roughly four times better neutralization activity than convalescent sera. Also in this study, ID and IM administration of mRNA-1273 resulted in considerably higher concentrations of antibodies than convalescent serum. Since ID vaccination induced antibody levels similar to IM administration, the antibody levels reported here likely have high neutralising capacity.

This study has some limitations. Although the concentrations of anti-S1 IgG were higher at each time point, the limited sample size did not allow to demonstrate superiority of intradermal delivery over intramuscular injection. In addition, since we included healthy volunteers aged 18-30 years old, the results on safety and immunogenicity may not apply to the general population. Finally, the longevity of the immune response past day 43 was not assessed, as some participants opted for receiving the regular vaccine through the national COVID-19 vaccination program at day 57.

Although the relative importance of neutralization antibodies with regard to protection from COVID-19 has not yet been fully characterized, high levels of neutralising antibodies have been shown in early preclinical Rhesus macaque studies^22^, in convalescent individuals^23,24^ and in recently reported vaccine trials.^25,26^ Additional measurements of neutralising antibodies and T-cell responses will be performed at a later part and will provide more information on the robustness and longevity of the immune response.

In conclusion, we show that the ID administration of 10 µg and 20 µg mRNA-1273 vaccine resulted in a robust, homogeneous, immune response with an acceptable safety profile in healthy adults aged 18-30 years. These safety and immunogenicity results support advancement of the investigation of the intradermal route for administration of the fractional doses of the mRNA-1273 vaccine to later-stage clinical trials. A randomised non-inferiority trial of 20 µg mRNA-1273 administered ID vs 100 µg mRNA-1273 administered IM is in preparation (part 3). When part 3 of this trial confirms these preliminary findings, ID administration will bear great potential as a dose-sparing strategy in countries where vaccination campaigns are still at an early stage and could be a true turning point in this pandemic that has gripped the world for one and a half years.

## Supporting information

Prespecified stopping rules

## Data Availability

Deidentified individual participant data will be made available immediately after publication and finalization of the completed clinical study report, upon requests directed to the corresponding author. After approval of proposal, data can be shared through a secure online platform.

