## Supplementary material for "Tolerability, safety and immunogenicity of intradermal delivery of a fractional dose mRNA-1273 SARS-CoV-2 vaccine in healthy adults as a dose sparing strategy": Figure S1

**DAY 1:**

NUMBER OF PARTICIPANT: IDSCOVA

##### |__|__|__|

| DATE DIARY:    \|__\|__\| \|__\|__\| \|__\|__\|__\|__\|   | TIME OF FILLING IN DIARY:    \|__\|__\| : \|__\|__\|   |
| --- | --- |

Please circle the answer that best applies to you or write a short comment:

| LOCAL REACTIONS: | Absent | Mild | Moderate | Severe | Potentially life threatening |
| --- | --- | --- | --- | --- | --- |
| Pain at injection site   | None | Does not interfere with daily activities | Interferes with daily activities | Severe pain | Emergency room visit or hospitalisation for severe pain |
| Muscle stiffness | None | Does not interfere with daily activities | Interferes with daily activities | Prevents daily activitie | Emergency room visit or hospitalisation for severe muscle stiffness |
| Redness   |   \|__\|__\|cm   |   \|__\|__\|cm   |   \|__\|__\|cm |   \|__\|__\|cm |   \|__\|__\|cm |
|  | <2.5 cm | 2.5 cm tot 5.0 cm | >5.0 cm tot 10 cm | > 10 cm | Necrosis or exfoliative dermatitis |
| Swelling   |   \|__\|__\|cm   |   \|__\|__\|cm   |   \|__\|__\|cm |   \|__\|__\|cm |   \|__\|__\|cm |
|  | <2.5 cm | 2.5 cm tot 5.0 cm | >5.0 cm tot 10 cm | > 10 cm | Necrosis |
|   Pain and swelling at regional lymph nodes | None | Does not interfere with daily activities | Interferes with daily activities | Prevents daily activities | Emergency room visit or hospitalisation for severe pain |
| SYSTEMIC REACTIONS: | Absent | Mild | Moderate | Severe | Potentially life threatening |
|   Vomiting | None | 1-2 times in 24 hours | >2 times in 24 hours | Requires IV hydration | Emergency room visit or hospitalisation for hyptensive shock |
|   Dhiarrhoea | None | 2 to 3 loose stools in 24 hours | 4 to 5 loose stools in 24 hours | 6 or more loose stools in 24 hours | Emergency room visit or hospitalisation for severe diarrhoea |
|   Headache | None | Does not interfere with daily activities | Some interference with daily activities | Prevents daily activities | Emergency room visit or hospitalisation for severe headache |
|   Fatigue/ tiredness | None | Does not interfere with daily activities | Some interference with daily activities | Prevents daily activities | Emergency room visit or hospitalisation for severe fatigue |
|   Chills | None |  Does not interfere with daily activities | Some interference with daily activities | Prevents daily activities | Emergency room visit or hospitalisation for severe chills |
|   New or worsened muscle pain | None | Does not interfere with daily activities | Some interference with daily activities | Prevents daily activities | Emergency room visit or hospitalisation for severe muscle pain |
|   New or worsened joint pain | None | Does not interfere with daily activities | Some interference with daily activities | Prevents daily activities | Emergency room visit or hospitalisation for severe joint pain |

| DAY 1: DATE DIARY    \|__\|__\| \|__\|__\| \|__\|__\|__\|__\|   | TIME OF FILLING IN DIARY:    \|__\|__\| : \|__\|__\|   |
| --- | --- |

PARTICIPANT NUMBER: IDSCOVA

##### |__|__|__|

| Temperature | Absent | Mild | Moderate | Severe | Very severe |
| --- | --- | --- | --- | --- | --- |
| \|__\|__\|__\| | <38°C | 38.0°C – 38.4°C | 38.5°C -38.9°C | 39.0°C -40.0°C | >40.0°C |

| New concomitant medication (date) | Name medication | Dose & units | How many times a day | Indication | By general practitioner? |
| --- | --- | --- | --- | --- | --- |
| from:  \|__\|__\| \|__\|__\| \|__\|__\|  to:  \|__\|__\| \|__\|__\| \|__\|__\| |  |   ______ | \|__\|__\|a day |  | \|__\| YES  \|__\| NO |
| from:  \|__\|__\| \|__\|__\| \|__\|__\|  to:  \|__\|__\| \|__\|__\| \|__\|__\| |  | ______ | \|__\|__\|a day |  | \|__\| YES  \|__\| NO |
| from:  \|__\|__\| \|__\|__\| \|__\|__\|  to:  \|__\|__\| \|__\|__\| \|__\|__\| |  | ______ | \|__\|__\|a day |  | \|__\| YES  \|__\| NO |
| from:  \|__\|__\| \|__\|__\| \|__\|__\|  to:  \|__\|__\| \|__\|__\| \|__\|__\| |  | ______ | \|__\|__\|a day |  | \|__\| YES  \|__\| NO |
| from:  \|__\|__\| \|__\|__\| \|__\|__\|  to:  \|__\|__\| \|__\|__\| \|__\|__\| |  | ______ | \|__\|__\|a day |  | \|__\| YES  \|__\| NO |
| from:  \|__\|__\| \|__\|__\| \|__\|__\|  to:  \|__\|__\| \|__\|__\| \|__\|__\| |  | ______ | \|__\|__\|a day |  | \|__\| YES  \|__\| NO |
| Other (systemic) reactions: | Remarks: | | | | |
