## Supplementary material for "Tolerability, safety and immunogenicity of intradermal delivery of a fractional dose mRNA-1273 SARS-CoV-2 vaccine in healthy adults as a dose sparing strategy": Prespecified stopping rules

The following stopping rules were in place for all participants, based on review of diary reactogenicity and AE data.

Stopping Rule Criteria Safety:

- If any participant vaccinated with mRNA-1273 (at any dose level) develops an SAE or SUSAR that is assessed by the investigator as possibly related, or for which there is no alternative, plausible, attributable cause.

- If any participant vaccinated with mRNA-1273 (at any dose level) develops a Grade 4 local reaction within 14 days after vaccination that is assessed as possibly related by the investigator, or for which there is no alternative, plausible, attributable cause.

- If any participant vaccinated with mRNA-1273 (at any dose level) develops a Grade 4 systemic event within 7 days after vaccination that is assessed as possibly related by the investigator, or for which there is no alternative, plausible, attributable cause

- If two participants vaccinated with mRNA-1273 (within the same dose level) develop a fever >40.0°C for at least one daily measurement within 7 days after vaccination that is assessed as possibly related by the investigator, or for which there is no alternative, plausible, attributable cause.

- If any two participants vaccinated with mRNA-1273 (at any dose level) report the same or similar severe (Grade 3) AE within 14 days after vaccination, assessed as possibly related by the investigator, or for which there is no alternative, plausible, attributable cause.

- If any participant dies or requires ICU admission due to SARS-CoV-2 infection; if this stopping rule is met, all available clinical and preclinical safety and immunogenicity data should be reviewed to evaluate for enhanced COVID-19 disease.

Stopping Rule Criteria Immunogenicity:

- If <12/15 participants vaccinated with mRNA-1273 (at 20 μg dose level) do not meet the threshold for neutralising antibodies (PRNT80 ≥128) at Day 43.

- If part 2 is unsafe and ≤7 out of 10 participants of part 1 developed virus neutralizing antibody titres (PRNT80) ≥128 at day 43,

The anti-Spike IgG antibody titres may be used as a proxy for the neutralizing antibody titres if the results of the neutralizing antibody assay are delayed.
