## Supplementary material for "Tolerability, safety and immunogenicity of intradermal delivery of a fractional dose mRNA-1273 SARS-CoV-2 vaccine in healthy adults as a dose sparing strategy": S1 tm S4

**Solicited adverse events**

Local reactions

*Redness and swelling* was measured and recorded in centimetres and categorised as absent, mild, moderate, or severe based on the grading scale (Table S1).

*Pain* at the injection site was assessed by the participant as absent, mild, moderate, or severe according the grading scale in Table S1.

Systemic events

Solicited systemic events consisted of vomiting, diarrhoea, headache, fatigue, chills, new or worsened muscle pain, and new or worsened joint pain. The symptoms were assessed by the participant as absent, mild, moderate, or severe according to the grading scale in Table S2.

Fever

Participants were instructed on how to measure oral temperature at home. Daily temperature measurements were registered in the diary in the morning and at any time when fever is suspected during the diary data collection periods. Fever was defined as an oral temperature of ≥38.0°C. The highest temperature for each day was recorded in the diary. Temperature was measured and recorded to 1 decimal place and then categorised during analysis according to the scale shown in Table S3.

**Table S1. Grading scale for severity of solicited vaccine-related local adverse reactions**

|  | **Mild (Grade 1)** | **Moderate (Grade 2)** | **Severe (Grade 3)** | **Potentially life threatening (Grade 4)** |
| --- | --- | --- | --- | --- |
| **Pain at injection site** | Does not interfere with activity | Interferes with activity | Prevents daily activity | Emergency room visit or hospitalization for severe pain |
| **Redness** | 2·5 cm to 5·0 cm | >5·0 cm to 10·0 cm | >10 cm | Necrosis or exfoliative dermatitis |
| **Swelling** | 2·5 cm to 5·0 cm | >5·0 cm to 10·0 cm | >10 cm | Necrosis |
| **Pain and swelling at regional lymph nodes** | Does not interfere with activity | Interferes with activity | Prevents daily activity | Emergency room visit or hospitalization for severe pain |

**Table S2. Grading scale for severity of solicited vaccine-related systemic adverse reactions**

|  | **Mild (Grade 1)** | **Moderate (Grade 2)** | **Severe (Grade 3)** | **Potentially life threatening (Grade 4)** |
| --- | --- | --- | --- | --- |
| **Vomiting** | 1-2 times in 24 hours | >2 times in 24 hours | Requires IV hydration | Emergency room visit or hospitalization for hypotensive shock |
| **Diarrhoea** | 2 to 3 loose stools in 24 hours | 4 to 5 loose stools in 24 hours | 6 or more loose stools in 24 hours | Emergency room visit or hospitalization for severe diarrhoea |
| **Headache** | Does not interfere with activity | Some interference with activity | Prevents daily routine activity | Emergency room visit or hospitalization for severe headache |
| **Fatigue/tiredness** | Does not interfere with activity | Some interference with activity | Prevents daily routine activity | Emergency room visit or hospitalization for severe fatigue |
| **Chills** | Does not interfere with activity | Some interference with activity | Prevents daily routine activity | Emergency room visit or hospitalization for severe chills |
| **New or worsened muscle pain** | Does not interfere with activity | Some interference with activity | Prevents daily routine activity | Emergency room visit or hospitalization for severe new or worsened muscle pain |
| **New or worsened joint pain** | Does not interfere with activity | Some interference with activity | Prevents daily routine activity | Emergency room visit or hospitalization for severe new or worsened joint pain |

**Table S3. Grading scale for severity of vaccine-related fever**

| **Absent** | <38·0°C |
| --- | --- |
| **Mild** | 38·0-38·4°C |
| **Moderate** | 38·5-38·9°C |
| **Severe** | 39·0-40·0°C |
| **Very severe** | >40·0°C |

**Table S4. Use of antipyretic medication for vaccine-related adverse events**

|  | **ID 10 µg** | **ID 20 µg** | **IM 20 µg** |
| --- | --- | --- | --- |
| **Vaccination 1** | 1/10 (10%) | 3/15 (20%) | 0/15 (0%) |
| **Vaccination 2** | 0 /9 (0%) | 3/15 (20%) | 1/14 (7.1%) |

Number of participants who used antipyretic medication after the first or second mRNA-1273 vaccination 10 μg or 20 µg intradermally (ID) or 20 μg intramuscularly (IM)
