## Supplementary material for "Tolerability, safety and immunogenicity of intradermal delivery of a fractional dose mRNA-1273 SARS-CoV-2 vaccine in healthy adults as a dose sparing strategy": Table S5

**Table S5. Duration of vaccine-related adverse events**

| **10 µg ID** | **Vaccination 1**  **n days (SD), range** | **Vaccination 2**  **n days (SD), range** |
| --- | --- | --- |
| *Local reactions* |  |  |
| Hyperpigmentation | 44 (-)  44-44 | 15 (1)  14-15 |
| Local muscle stiffness | 1 (0)  1-1 | ·· |
| Itch at injection site | 2 (1)  1-2 | 4 (3)  1-7 |
| Pain at injection site | 3 (1)  2-4 | 3 (1)  2-4 |
| Swelling | 2 (-)  2-2 | 2 (-)  2-2 |
| Erythema | 2 (1)  1-2 | 2 (1)  1-3 |
| *Systemic reactions* |  |  |
| Headache | 1 (0)  1-1 | ·· |
| **20 µg ID** | **Vaccination 1**  **n days (SD), range** | **Vaccination 2**  **n days (SD) , range** |
| *Local reactions* |  |  |
| Local muscle stiffness | 2 (1)  1-4 | 2 (1)  1-6 |
| Itch at injection site | 3 (2)  1-7 | 3 (2)  1-8 |
| Pain at injection site | 2 (1)  1-4 | 3 (1)  1-5 |
| Swelling | 2 (1)  1-4 | 3 (1)  2-6 |
| Erythema | 2 (1)  1-6 | 3 (1)  1-6 |
| Axillary lymphadenopathy | ·· | 2 (0)  2-2 |
| *Systemic reactions* |  |  |
| Emesis | ·· | 1 (-)  1-1 |
| Myalgia | 2 (1)  1-3 | 1 (1)  1-2 |
| Headache | 1 (0)  1-1 | 2 (1)  1-2 |
| Diarrhoea | ·· | ·· |
| Chills | 1 (-)  1-1 | 1 (-)  1-1 |
| Fatigue | 2 (1)  1-4 | 2 (1)  1-3 |
| **20 µg IM** | **Vaccination 1**  **n days (SD) , range** | **Vaccination 2**  **n days (SD) , range** |
| *Local reactions* |  |  |
| Local muscle stiffness | 2 (1)  1-3 | 2 (1)  1-3 |
| Pain at injection site | 2 (1)  1-3 | ·· |
| Swelling | 1 (-)  1-1 | 2 (1)  1-2 |
| Erythema | 1 (-)  1-1 | 2 (1)  1-2 |
| *Systemic reactions* |  |  |
| Myalgia | 2 (-)  2-2 | 1 (1)  1-2 |
| Headache | 1 (1)  1-1 | 1 (0)  1-2 |
| Diarrhoea | 1 (-)  1-1 | 2 (1)  1-4 |
| Chills | 1 (-)  1-1 | 1 (0)  1-1 |
| Fatigue | 2 (1)  1-2 | 2 (1)  1-3 |
| Athralgia | ·· | 1 (-)  1-1 |
