## Supplementary material for "Tolerability, safety and immunogenicity of intradermal delivery of a fractional dose mRNA-1273 SARS-CoV-2 vaccine in healthy adults as a dose sparing strategy": Table S6

**Table S6. Characteristics of selected participants of PIENTER-Corona with mild to moderate COVID-19 or mRNA-1273 vaccination**

|  | **Mean** | **95% CI** |  | **N** | **%** |
| --- | --- | --- | --- | --- | --- |
|  |  | **lower** | **upper** |  |  |
| **COVID-19 patients** |  |  |  |  |  |
| Age | 23 | 27·0 | 32·76 |  |  |
| Sex (female) |  |  |  | 17 | 73·9 |
| Days since symptoms | 41 | 34·8 | 46·5 |  |  |
| Medical consultation |  |  |  | 4 | 17·4 |
| Fatigue |  |  |  | 6 | 26·1 |
| Ageusia/Anosmia |  |  |  | 36 | 67·9 |
| Headache |  |  |  | 12 | 52·2 |
| Myalgia |  |  |  | 12 | 52·2 |
| Fever |  |  |  | 12 | 52·2 |
| Coughing |  |  |  | 16 | 69·6 |
| Dyspnea |  |  |  | 3 | 13·0 |
| **Vaccinated individuals** |  |  |  |  |  |
| Age | 44 | 39·2 | 49·57 |  |  |
| Sex (female) |  |  |  | 19 | 95·0 |
| Days since second vaccination | 38·8 | 33·8 | 43·8 |  |  |

Characteristics of PCR-confirmed patients with mild to moderate COVID-19 of whom serum was collected between two weeks and two months after symptom onset, and of 20 vaccinated individuals without an infection history, aged 18-65 years, collected between 14 and 45 days after receiving two doses of 100 µg mRNA-1273 vaccine IM
