## Supplementary material for "Tolerability, safety and immunogenicity of intradermal delivery of a fractional dose mRNA-1273 SARS-CoV-2 vaccine in healthy adults as a dose sparing strategy": Figure S2

**Figure S2. Photographs of local adverse events**

| **A** | 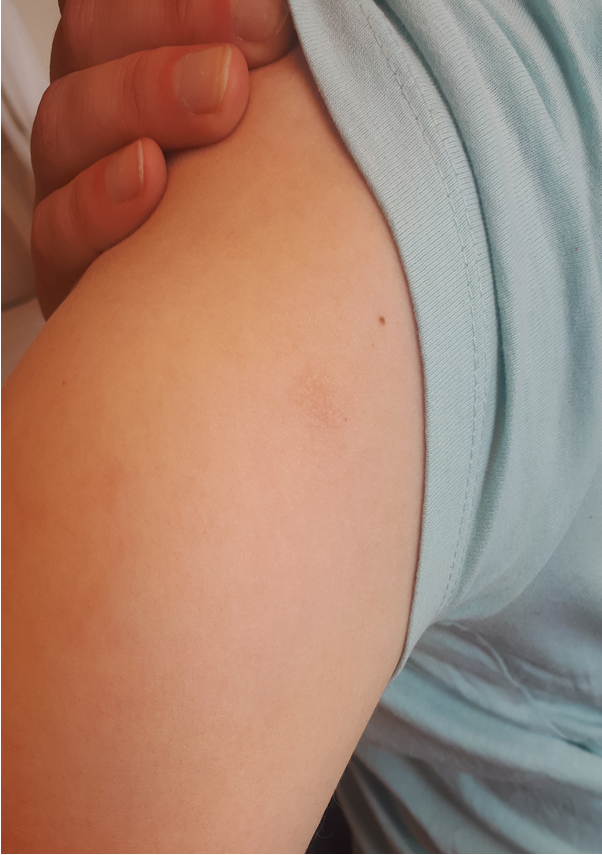 |
| --- | --- |
| **B** | 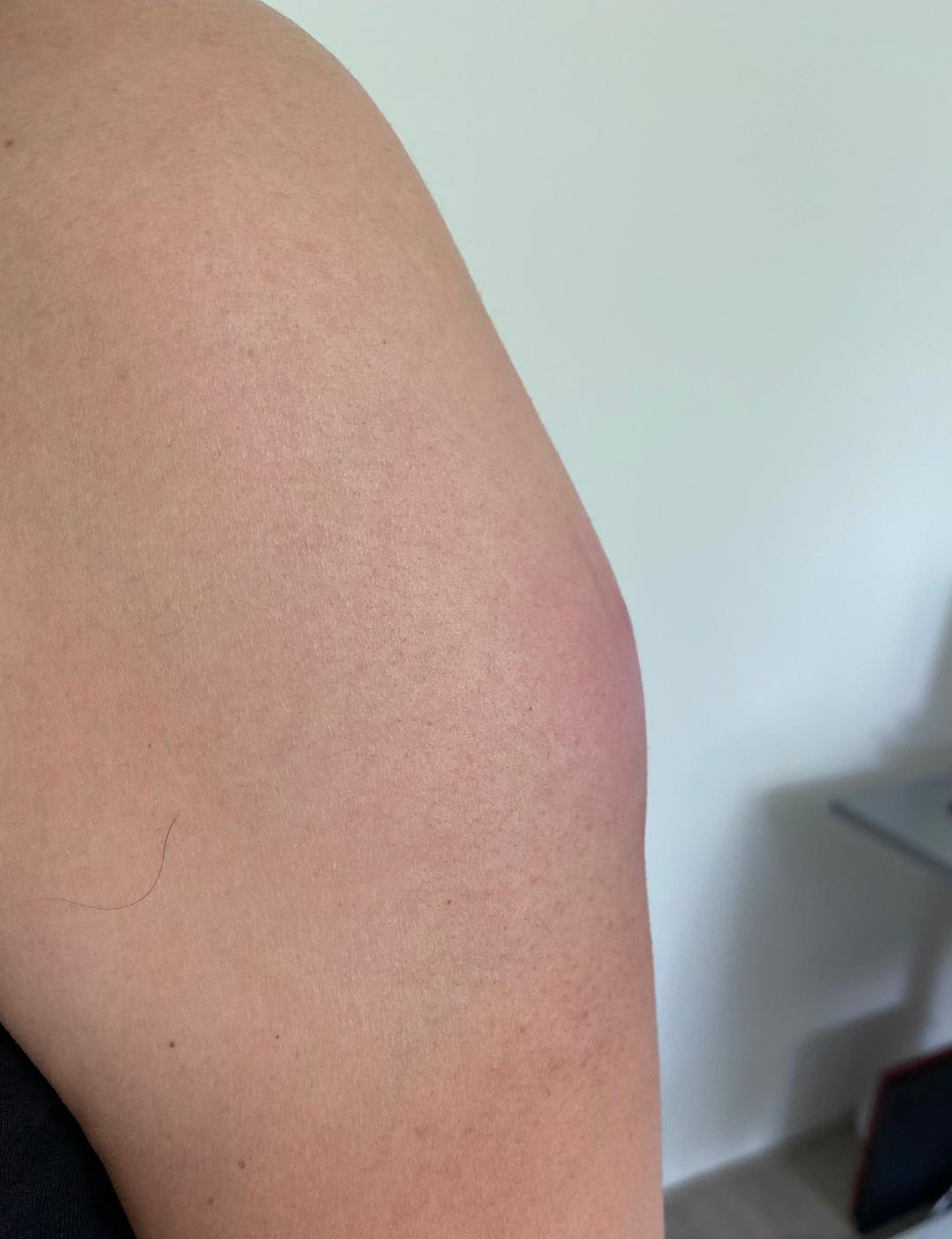 |
| **C** | 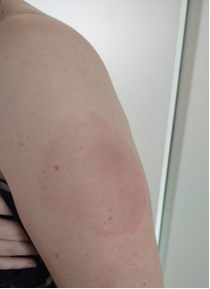 |
| **D** | 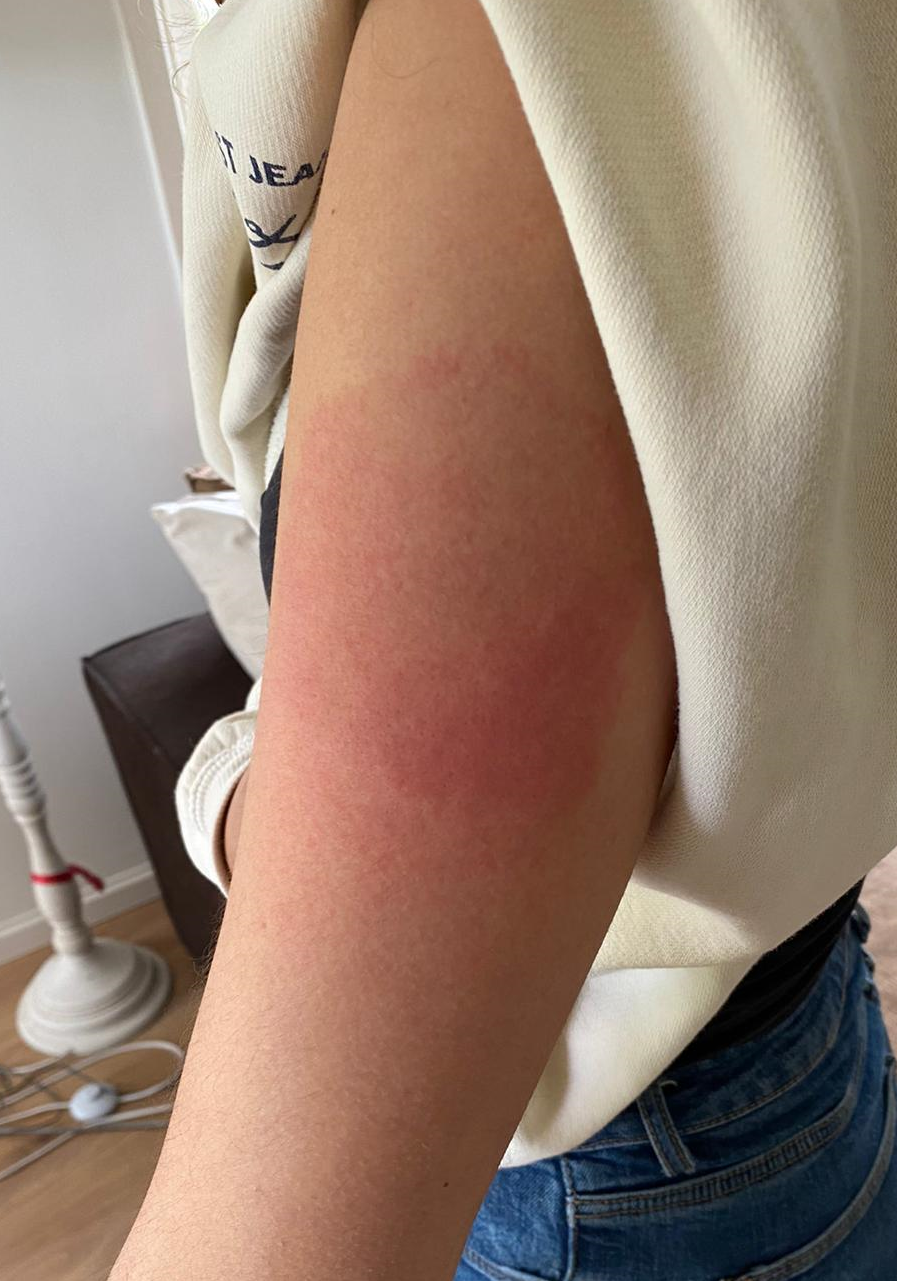 |
| Photographs of local adverse events: (A) mild hyperpigmentation after first vaccination with 10 µg ID; (B) moderate swelling and erythema in 20 µg ID group; (C) severe erythema in 20 µg ID group; (D) moderate erythema and swelling in 20 µg IM group. Photographs are published with permission of participants | |
