## Supplementary material for "Tolerability, safety and immunogenicity of intradermal delivery of a fractional dose mRNA-1273 SARS-CoV-2 vaccine in healthy adults as a dose sparing strategy": Figure S3

**Figure S3*.* Anti-spike S1 IgG antibody concentrations.**

**A**

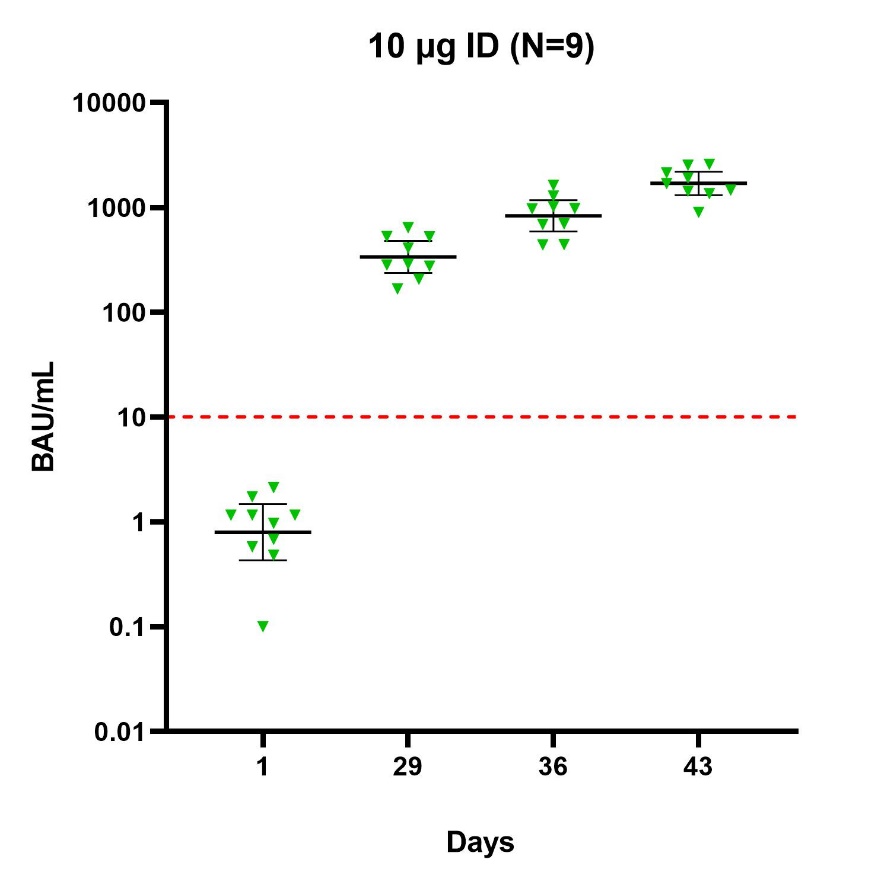

**B**

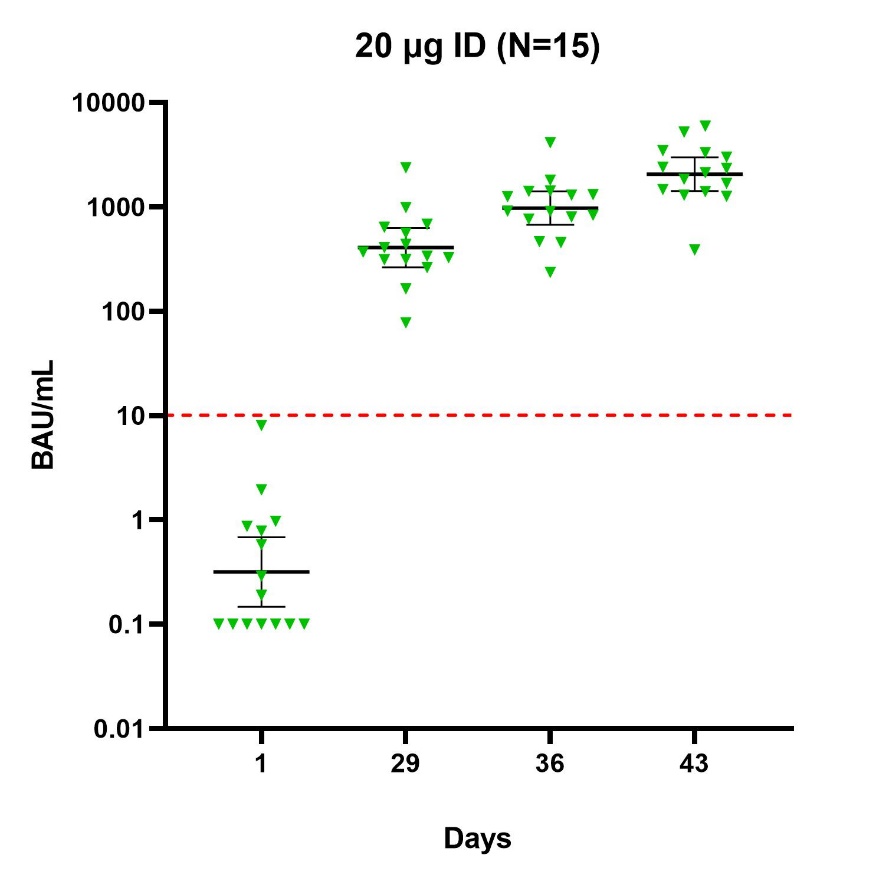

**C**

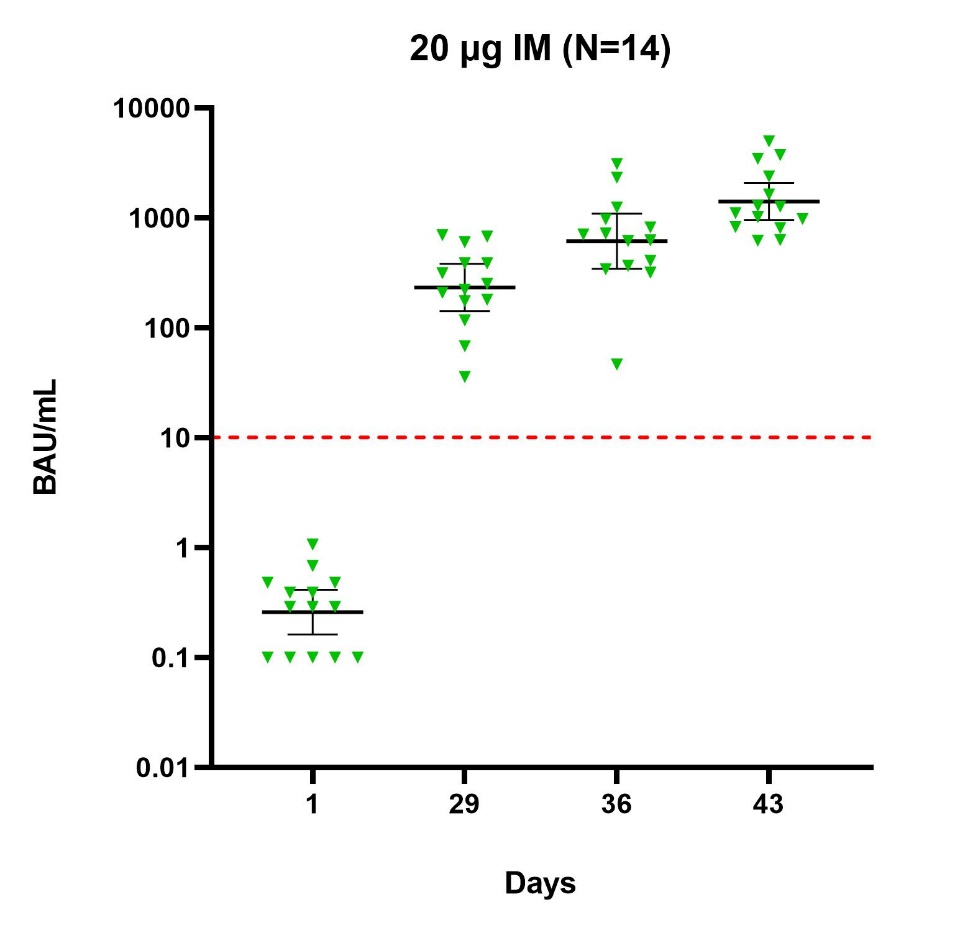

Anti-Spike S1 IgG levels reported in binding antibody units/mL at day 1, 29, 36 and 43 after (A) 10 μg ID; (B) 20 μg ID or (C) 20 µg IM. Each symbol represents a sample from an individual participant. Lines indicate GMC and 95% confidence intervals. Cut-off for seropositivity is 10·08 (dotted line).
