## Supplementary material for "Tolerability, safety and immunogenicity of intradermal delivery of a fractional dose mRNA-1273 SARS-CoV-2 vaccine in healthy adults as a dose sparing strategy": Figure S5

**Figure S5. Average diameter of swelling and erythema after vaccination**

**A**

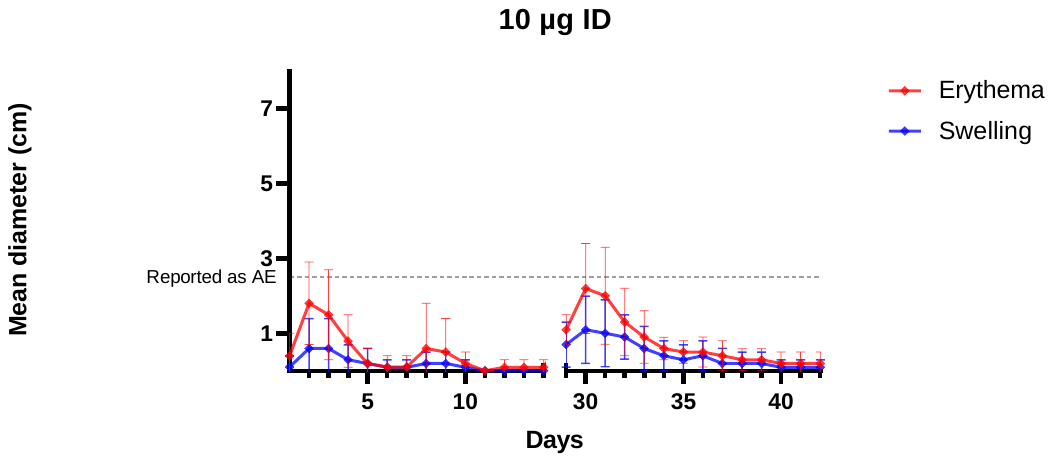

**B**

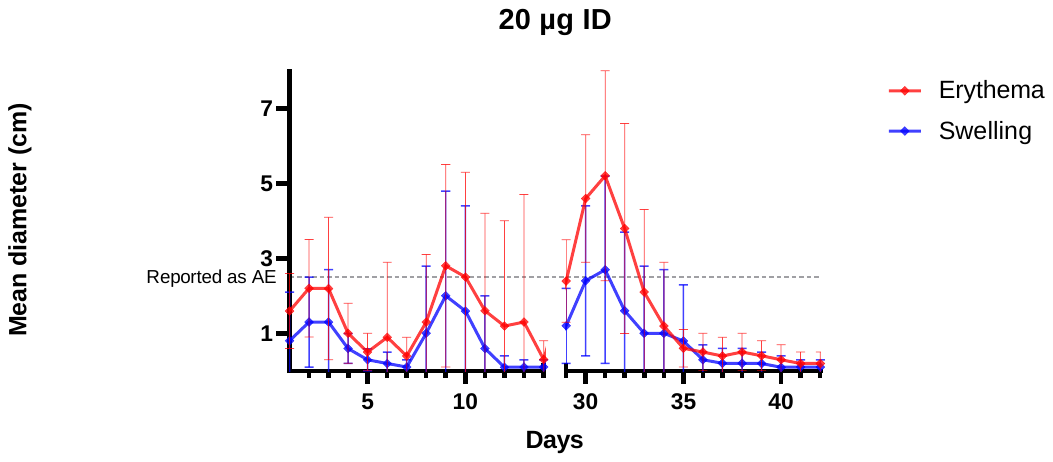

**C**

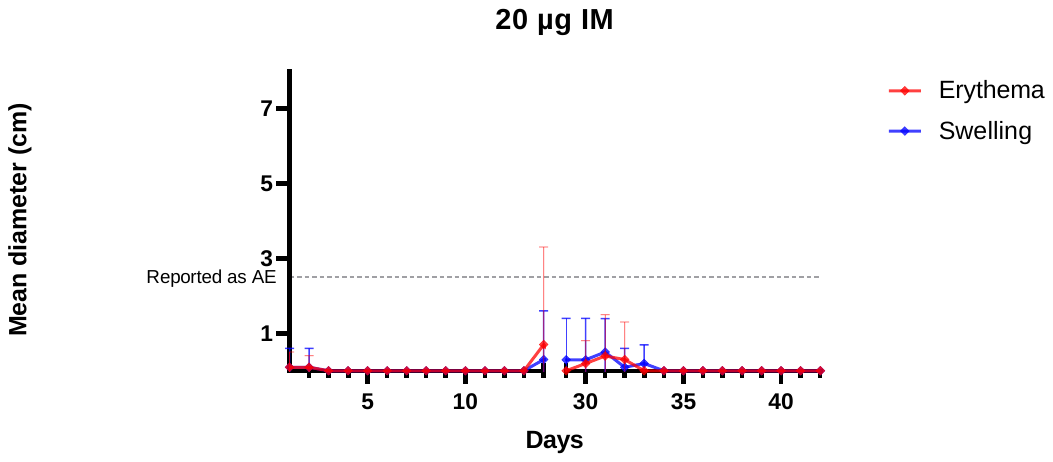

Average diameter of local swelling and erythema after vaccination on day 1, and day 29 with (A) 10 μg ID; (B) 20 μg ID or (C) 20 µg IM. Error bars represent standard deviation. Swelling and erythema of ≥ 2.5 cm was reported as separate adverse event (dotted line).
